# Nanopore-based hybridization capture approaches for targeted viral metagenomics and whole-genome sequencing

**DOI:** 10.1101/2025.04.08.25325453

**Authors:** Oscar Enrique Torres Montaguth, Sarah Buddle, Leysa Forrest, Tony Brooks, Helena Tutill, Peter Simmonds, Heli Harvala, Tanya Golubchik, Rachel Williams, Sofia Morfopoulou, Judith Breuer

**Author notes:** These authors contributed equally (Oscar Enrique Torres Montaguth, Sarah Buddle).

## Abstract

Hybridization capture approaches can significantly increase the sensitivity of metagenomics. However, their implementation with Oxford Nanopore Technologies (ONT) sequencing has been limited due to the lack of dedicated workflows and the need for an extra ligation-based library preparation step. We developed a universal four-primer PCR approach that allows the implementation of hybridization capture workflows with ONT without additional ligation-based steps. Our method allowed the fast conversion of a targeted metagenomics workflow, increasing ONT sensitivity by 10 to 100-fold compared to untargeted approaches and providing both rapid detection and whole-genome sequences where the pathogen was abundant and identification where viral loads were low.

## Background

Metagenomics, the comprehensive sequencing of all the genetic material present in a sample, has the potential to become a single-test method for the unbiased detection of known and novel viruses. This makes it a powerful alternative to traditional methods, not only for diagnosis of infection but also for surveillance of emerging viruses and pandemic preparedness [1–3]. A range of metagenomics workflows have been evaluated and implemented in different clinical settings, particularly for the diagnosis of complex clinical cases like encephalitis and respiratory tract infections (RTIs) [4–8], and more recently for syndromic and wastewater-based surveillance [9–11].

Widespread adoption of metagenomics has been limited by high equipment and reagent costs, complex laboratory and data analysis workflows, long turnaround times, and reduced sensitivity compared to other molecular methods like qPCR [2]. Most metagenomics protocols use short read sequencing platforms such as Illumina and have turnaround times of days to weeks and high equipment costs. Oxford Nanopore Technologies (ONT) sequencing allows the development of rapid sequencing workflows by enabling real time data analysis, and due to its reduced footprint and equipment costs, it can be easily implemented in diverse clinical settings [12–15]. Despite these advantages, sensitivity for viruses can be limited, particularly in samples with low viral loads or high levels of host nucleic acids [16,17]. Depletion of background host material can improve detection of DNA pathogens and to a lesser extent RNA viruses but may deplete intracellular pathogens and cell-free DNA [2,12,15,18].

Hybridization capture methods can significantly increase sequencing sensitivity compared to untargeted approaches. These protocols use biotinylated RNA or DNA oligonucleotides that hybridize to targets of interest, enriching them for sequencing. Hybridization capture workflows can provide an increase of 10 to 100-fold when compared with untargeted approaches [17,19,20]. Probes can efficiently enrich for targets with 20% difference to the reference sequence used for probe design, allowing the detection of divergent pathogens [21,22]. Comprehensive panels have been designed and used for the improved detection of viruses, bacteria and eukaryotic pathogens in diverse sample types [16,19,20,23–26]. However, hybridization capture workflows are long and complex, and have been used mostly in combination with short-read sequencers, leading to long turnaround times and higher costs, thus limiting their application in clinical settings.

Implementation of hybridization capture methods in ONT-based workflows has been limited, since available commercial protocols are designed for short-read platforms. Current approaches to adapt existing workflows for ONT require an additional ligation-based library preparation step at the end of the workflow [27–29]. This second step adds around 1.5 to 3 hours, depending on sample number and whether an extra ONT barcoding step is performed, increasing the complexity of the workflow and leading to longer turnaround times. Additionally, end-repair and ligation accessory kits are required, increasing the processing costs per sample.

Here we present a fast universal four-primer PCR method that allows the use of existing hybridization capture workflows with ONT without the need for extra ligation-based library preparation steps. We evaluated the performance of our protocol in two applications: targeted metagenomics and whole-genome sequencing (WGS) of viruses. For targeted viral metagenomics, we tested the Twist Bioscience Comprehensive Viral Research Panel (CVRP) on samples consisting of a dilution series of a six-virus mock community in a human DNA/RNA background. We performed real-time data analysis and evaluated viral detection and genome coverage for the six members of the viral community. Additionally, we established the sensitivity, specificity and limits of detection of the ONT-CVRP method and compared its performance with untargeted ONT metagenomics. For viral WGS, we tested the Agilent SureSelect XT capture protocol using mock DNA and RNA samples spiked with human betaherpesvirus 5 DNA and SARS-CoV-2 RNA.

## Methods

### Mock clinical samples

We tested the performance of the Twist CVRP combined with ONT to evaluate the conversion method. We used simulated clinical samples containing a range of viruses and a high human nucleic acid content (40 ng/µl) which were produced and sequenced in a previous study [17]. They consisted of the ATCC Virome Virus mix (ATCC, MSA-1008) in four concentrations (60,000, 6,000, 600 and 60 genome copies per ml (gc/ml)) designed to match typical viral loads found in clinical samples, combined with commercially available DNA (Promega, G1471) and RNA (Invitrogen, AM7962). Lambda phage DNA (Sigma, D3779) and MS2 phage RNA (Roche, 10165948001) were used as internal controls in all samples. Two external control samples, consisting of the human nucleic acid mix and the positive control phages, but without the mock community viruses, were run in parallel, and two replicates were performed for the 600 gc/ml and 60 gc/ml samples, bringing the total number of samples to eight.

For WGS we used simulated clinical samples with viral loads compatible with the SureSelect XT workflow [30]. For human betaherpesvirus 5 (ATCC, VR-538DQ), we used a human DNA content expected for a blood sample (40ng/µl) spiked with4 x 10^4^ human betaherpesvirus 5 genome copies per microliter (gc/µl). For SARS-CoV-2 (NIBSC,101029) we used a simulated sample with a human RNA content expected for a nasopharyngeal swab (5 ng/µl) with a concentration of 2 x 10^4^ SARS-CoV-2 gc/µl.

### Targeted ONT viral metagenomic sequencing with Twist Comprehensive Viral Research Panel

Combined DNA/RNA hybridization capture was performed using the Twist Comprehensive Viral Research Panel (Twist Bioscience, 103550). Library preparation and pre-capture PCR was performed following the “Twist Bioscience Total Nucleic Acids Library Preparation EF Kit 2.0 for Viral Pathogen Detection and Characterization” protocol. Firstly, cDNA synthesis was performed using ProtoScript II First strand synthesis kit (New England Biolabs, E6560) followed by the NEBNext Ultra Non-Directional Second Strand Synthesis module (New England Biolabs, E6111). A total of 25ng of the double-stranded (ds) cDNA and dsDNA mix were used as input for adaptor ligation, indexing and pre-capture amplification using the Twist Library preparation EF Kit 2.0 (Twist Bioscience, 104207 and 100573).

Following pre-capture amplification, enrichment and washing was performed according to the “Twist Target Enrichment standard hybridization V1” protocol until just before post-capture PCR. Indexed samples were pooled, for a total of six samples plus two negative controls per hybridisation reaction, making a total of eight samples per reaction.

Hybridisation was performed overnight for 16 hours. Hybridisation targets were then captured with Streptavidin Binding Beads. At this step, samples were washed using the Twist Wash Buffers v1 kit (Twist Bioscience, 104178) and the streptavidin beads were resuspended in 45 µl of nuclease-free water (streptavidin bead slurry).

For post-capture PCR we used a four-primer PCR that allows the generation of a library compatible with ONT rapid sequencing adaptors. A PCR premix was prepared using 25 µl of NEBNext Ultra II Q5 Master mix (New England Biolabs, M0544), 1 µl of 2 µM (each) P-RPB primer mix (P5-RPB, 5’-CGTTTTTCGTGCGCCGCTTCAATGATACGGCGACCA CCGA -3’; P7-RPB, 5’-CGTTTTTCGTGCGCCGCTTCCAAGCAGAAGACGGCATACGA -3’) and 1.5 µl of Rapid Barcode Primer (RLB, Oxford Nanopore Technologies, SQK-RPB114.24). 22.5 µl of the streptavidin bead slurry were added to the reaction and mixed gently by pipetting until beads were fully resuspended. The reaction was placed immediately on a pre-heated PCR machine at 98°C and the following program was started: 98°C for 30 seconds; 5 cycles of 98°C for 10 seconds, 65°C for 20 seconds and 72°C for 20 seconds; 30 cycles of 98°C for 15 seconds, 71°C for 20 seconds and 72°C for 20 seconds followed by a final extension step of 72°C for 2 minutes. After PCR, the supernatant was recovered and DNA purified using a 0.8X volume ratio of Ampure beads following the Rapid Barcoding PCR kit 24 V14 instructions (Oxford Nanopore Technologies, SQK-RPB114.24). After purification, 125 fmol of the library were used for Rapid Adaptor (RAP) attachment. Libraries were run for 72 hrs on PromethION flow cells (Oxford Nanopore Technologies, FLO-PRO114M) on a P2 Solo connected to a GridION.

### Data preprocessing

Data was basecalled, demultiplexed and trimmed using dorado v0.9.0 with HAC and SUP and dual-end and single-end barcoding. Demultiplexing and trimming was performed first with the TWIST-96A-UDI kit option and then an additional trimming step using custom primer sequences was performed. The HAC dual-end data was used for analysis unless stated otherwise.

### Viral detection and coverage over time

Classification was performed using EPI2ME wf-metagenomics [31] with a custom database (see taxonomic classification section), which can be performed in real time. To evaluate viral detection and genome coverage over time, read timing data was extracted from the sequencing summary file, reads classified by EPI2ME (Kraken2 output) were aligned to the viral reference genomes using minimap2 [32] and coverage was calculated using samtools [33].

### Comparison with untargeted ONT

Previously published untargeted ONT datasets that were generated from the same mock clinical samples [17] were used for comparison. In the previous study, ten DNA samples were run on one PromethION flow cell and nine RNA on another. For this study, subsamples of these basecalled datasets were taken as if eight DNA and eight RNA samples were sequenced on the same flow cell to enable a comparison with the sequencing of eight combined DNA+RNA samples per flow cell with the Twist CVRP.

Reads preprocessed (trimmed and human filtered) through the nf-core Taxprofiler pipeline [34] were aligned to the reference genomes using minimap2 [32]. Mapped reads were extracted, PCR duplicates were removed, and coverage was calculated using samtools [33]. For the untargeted ONT, alignment of the DNA-Seq data is shown for the DNA viruses and RNA-Seq results for the RNA viruses.

### Taxonomic classification

In addition to EPI2ME, the trimmed outputs of dorado were classified using Kraken2 [35] run through the nf-core Taxprofiler pipeline [34]. Bracken [36] was performed on the Kraken2 outputs as a separate step. The same custom database was used for EPI2ME, Kraken2 and Bracken, consisting of all the bacterial (complete only), viral, fungal and protozoal genomes in RefSeq as of June 2023, with unplaced contigs removed [17,37]. The reads were also classified using CZID, a freely available online tool for metagenomics analysis, which has its own internal database bases on the NCBI NT and NR databases [38]. The nt_count field was used for analysis. All tools were run with the default parameters. Filters were applied using the metathresholds package using previously established thresholds (bacteria, fungi, other eukaryotes: reads per million ratio > 10 & proportion microbial reads > 1%, viruses: reads per million ratio > 5 or no reads in negative control & proportion microbial reads > 0.01%) [17,39]. For untargeted ONT data, data from DNA and RNA were combined for species counts, using the higher result for analysis.

### Targeted whole genome sequencing using SureSelect XT HS

Viral WGS was performed using SureSelect XT Low Input (Agilent Technologies, G9703A) in combination with probe panels for human betaherpesvirus 5 (Agilent Technologies, 5191-6707) and SARS-CoV-2 (Agilent Technologies, 5191-6838) following manufacturer instructions. For SARS-CoV-2 ds cDNA synthesis was performed using SuperScript IV RT (ThermoFisher Scientific, 18090200) for first-strand synthesis followed by a second-strand synthesis step using the NEBNext Ultra™ II Non-Directional RNA Second Strand Synthesis Module (New England Biolabs, E6111). DNA and dscDNA were fragmented using 8 microTUBE-50 AFA Fiber H Slit Strips (Covaris, 520243) on a Covaris E220 Focused-ultrasonicator prior to library preparation. 200 ng of DNA (human betaherpesvirus 5) and 120 ng of cDNA (SARS-CoV-s) were used as input for WGS. Library preparation, pre-capture PCR and hybridization capture were performed according to the “SureSelect XT Low input” protocol until just before post-capture PCR.

Similarly to the Twist CVRP panel, we used our four-primer PCR to generate libraries compatible with ONT rapid sequencing adaptors. A PCR premix was prepared using 25 µl of NEBNext Ultra II Q5 Master mix (New England Biolabs, M0544), 1 µl of 2 µM P-RPB primer mix, 1.5 µl of Rapid Barcode Primer and 10 µl of nuclease free water. 12.5 µl of the streptavidin bead slurry were added to the reaction and mixed gently by pipetting until beads were fully resuspended. Reaction was performed using the same PCR conditions described for the Twist CVRP workflow. After PCR, supernatant was recovered and DNA purified using a 0.8X volume ratio of Ampure beads following the Rapid Barcoding PCR kit 24 V14 instructions (Oxford Nanopore Technologies, SQK-RPB114.24). After purification, 100 fmol of the library were used for Rapid Adaptor (RAP) attachment. Libraries were run on MinION flow cells (Oxford Nanopore Technologies, FLO-MIN114) on a GridION.

### WGS data analysis

Data was basecalled with MinKnow [40] using HAC and demultiplexed and trimmed with dorado v0.9.0, using the SQK-RPB114-24 barcodes [41]. An additional trimming step was performed using custom primer sequences. The reads were aligned to the reference genome sequences with minimap2 [32] and aligned reads were extracted, duplicates removed and depth and coverage calculated using samtools [33]. Alignment plots were produced from samtools depth outputs using a custom R script.

### Code availability

Scripts used for analysis can be found at https://github.com/sarah-buddle/targeted-ont.

### Plots

Plots were produced in R using Tidyverse [42] packages or using Biorender.com.

## Results

### Targeted metagenomics

#### Optimizing a four-primer PCR approach for ONT conversion

To develop a fast conversion method for the implementation of hybridization capture methods in ONT workflows, we used a rapid sequencing approach that uses fast click chemistry to attach sequencing adaptors to the library. To achieve this, we developed a four-primer PCR that generates products compatible with ONT’s rapid adaptor protocol (**Figure 1A**). The four-primer PCR approach involves two PCR cycles in the same reaction. During the first stage of the reaction, primers P5-RPB and P7-RPB bind to primer binding sites (P5 and P7) previously introduced during pre-capture indexing PCR. The P5-RPB and P7-RPB primers add an RPB primer binding site to the library. In the second part of the reaction, ONT RLB primers from the Rapid PCR Barcoding kit bind to the RPB binding site introduced in the first part of the reaction, allowing for the introduction of ends compatible with ONT rapid adaptors during the post-capture library amplification step. After clean-up of the amplified library, sequencing adaptors (RAP) are attached by click chemistry and libraries are ready for sequencing.

**Figure 1:**
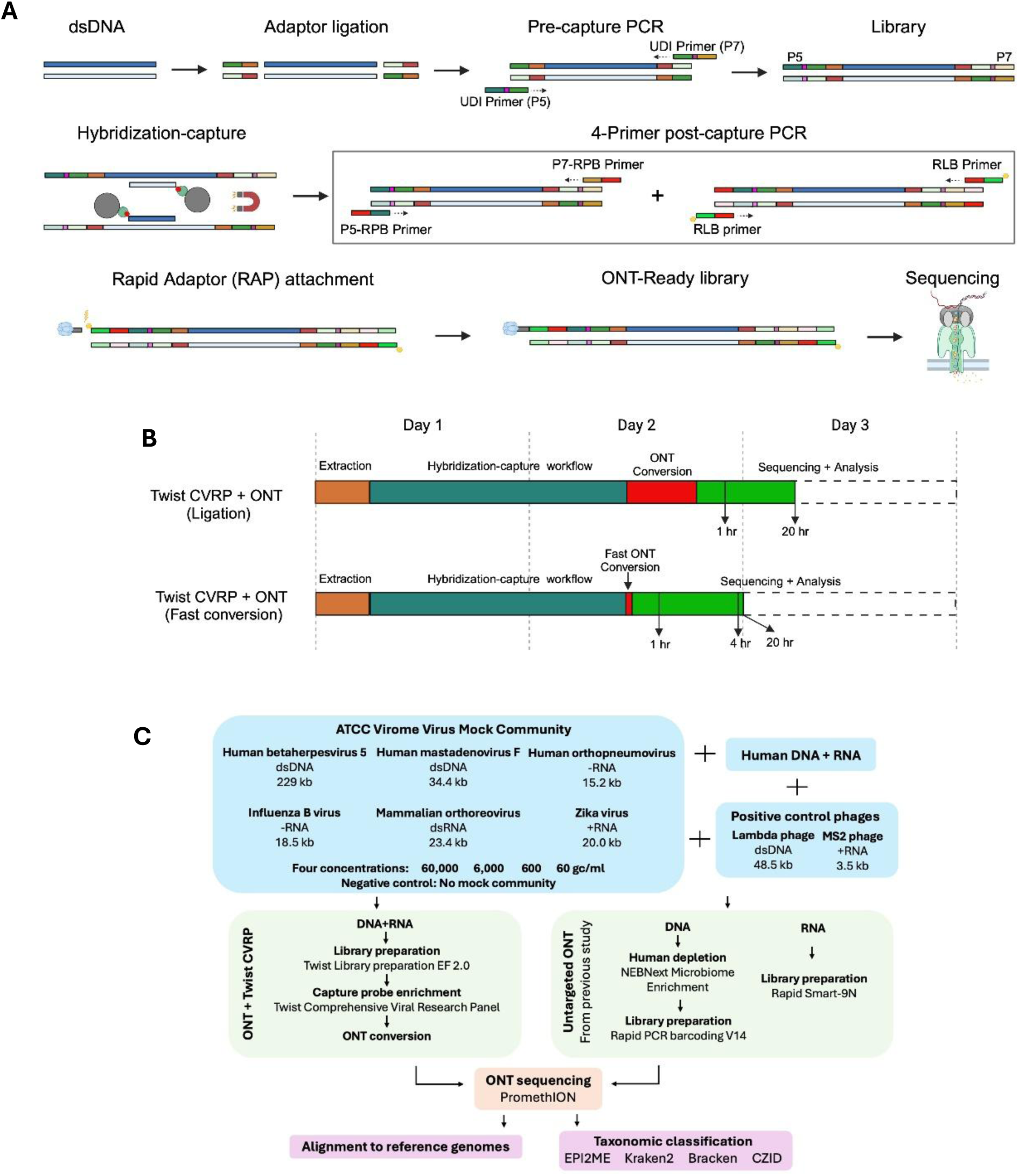
ONT hybridization capture workflow and experimental outline. A: ONT hybridization capture workflow using our four-primer PCR method. B: ONT hybridization capture turnaround times using a ligation-based approach and our four-primer PCR method. Potential reporting times after 1, 4 and 20 hours of the start of the sequencing run are indicated. C: Experimental outline for ONT metagenomics comparison. Created using biorender.com.

By implementing our four-primer PCR, conversion time can be reduced from 2-3 hours (depending on the number of samples) to approximately 15 minutes. The Twist CVRP workflow takes approximately 1.5 days, including an overnight hybridization step. By reducing the time spent on the conversion step, more sequencing data can be acquired and analyzed during the second day of the protocol, potentially providing early virus detection (**Figure 1B**).

#### Sequencing outputs

We tested the method on simulated clinical samples produced previously [17], which consist of a six-virus (two DNA and four RNA) mock community in four different concentrations in a human DNA/RNA background (**Figure 1C**). Samples were processed using the standard Twist CVRP protocol but the post-capture PCR step was replaced by the four-primer PCR, generating libraries ready for sequencing upon rapid adaptor attachment. Eight samples were sequenced together on a PromethION flow cell. This resulted in a total of 69.6 million basecalled reads (N50 = 450 bp), with 2.1-4.0 million reads and 0.3-0.6 Gb of usable sequence data per sample following demultiplexing and trimming (N50 = 160 bp) (**Table 1, Table S1**). A set of duplicate samples were run on a second flow cell, indicating how increasing sequencing depth by using more flow cells or decreasing the number of samples per flow cell could improve detection and coverage (**Table S2**).The positive control phages were not targeted by the TWIST CVRP panel and were not reliably present in the generated datasets (MS2 phage detected by alignment in 0/8 and lambda phage in 6/8 samples), demonstrating the need for alternative positive controls in future (**Table S3**). We proceeded with analysis, as the mock community viruses were detected in most samples, and at least 2 million reads were generated per sample **(Table 1)**.

**Table 1:**
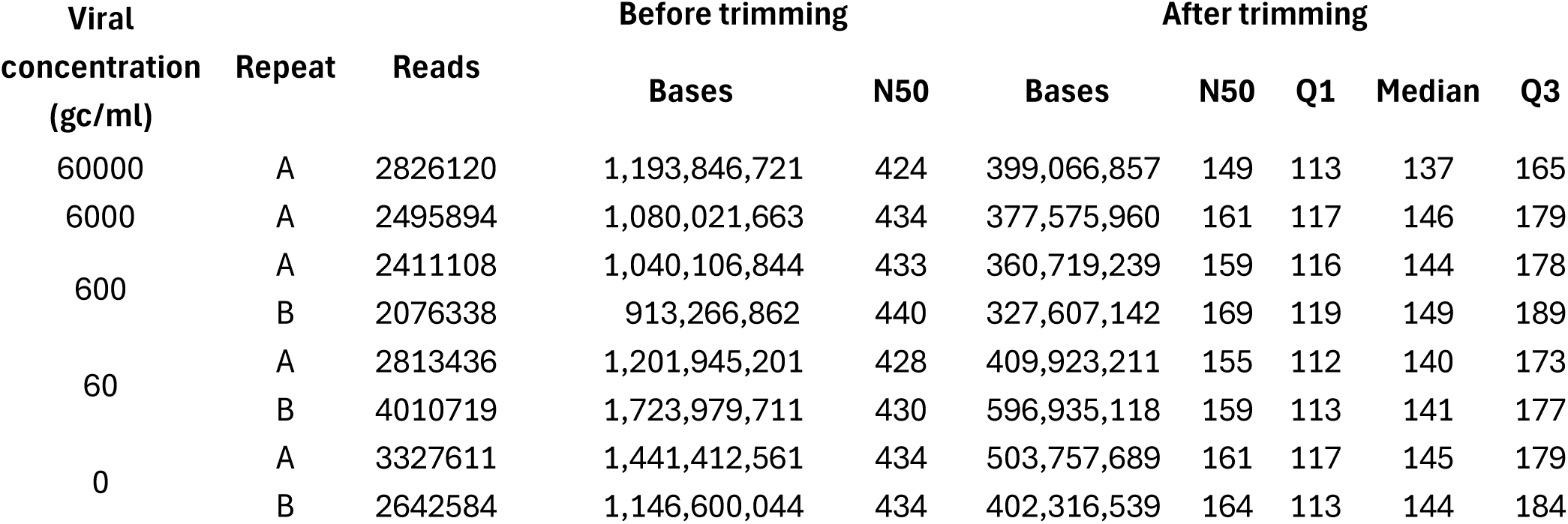
Sequencing output at 72 hours before and after adaptor trimming.

#### Detection and genome coverage of viruses

Data was analysed with the ONT-developed bioinformatic tools dorado [41] and EPI2ME [43], which can be used in real time. Using the data from one flow cell, we detected all six viruses at 60,000 gc/ml after 30 minutes of sequencing (**Figure 2A**) and detection of all viruses at 6,000 gc/ml within 2 hours (**Figure 2B**). Six and four out of six viruses were detected at 600 gc/ml and 60 gc/ml respectively by the end of the sequencing run at 72 hours (**Figure 2C&D)**. Near-full (>98%) genomes were obtained for Zika virus and mammalian orthoreovirus (genome sizes 20.0 kb and 23.4 kb respectively) at 60,000 gc/ml by 5h20m and 3h30m respectively (**Figure 3**). Genome coverage was at least 71% for all the viruses at 60,000 gc/ml by the end of the run (**Figure 3**).

**Figure 2:**
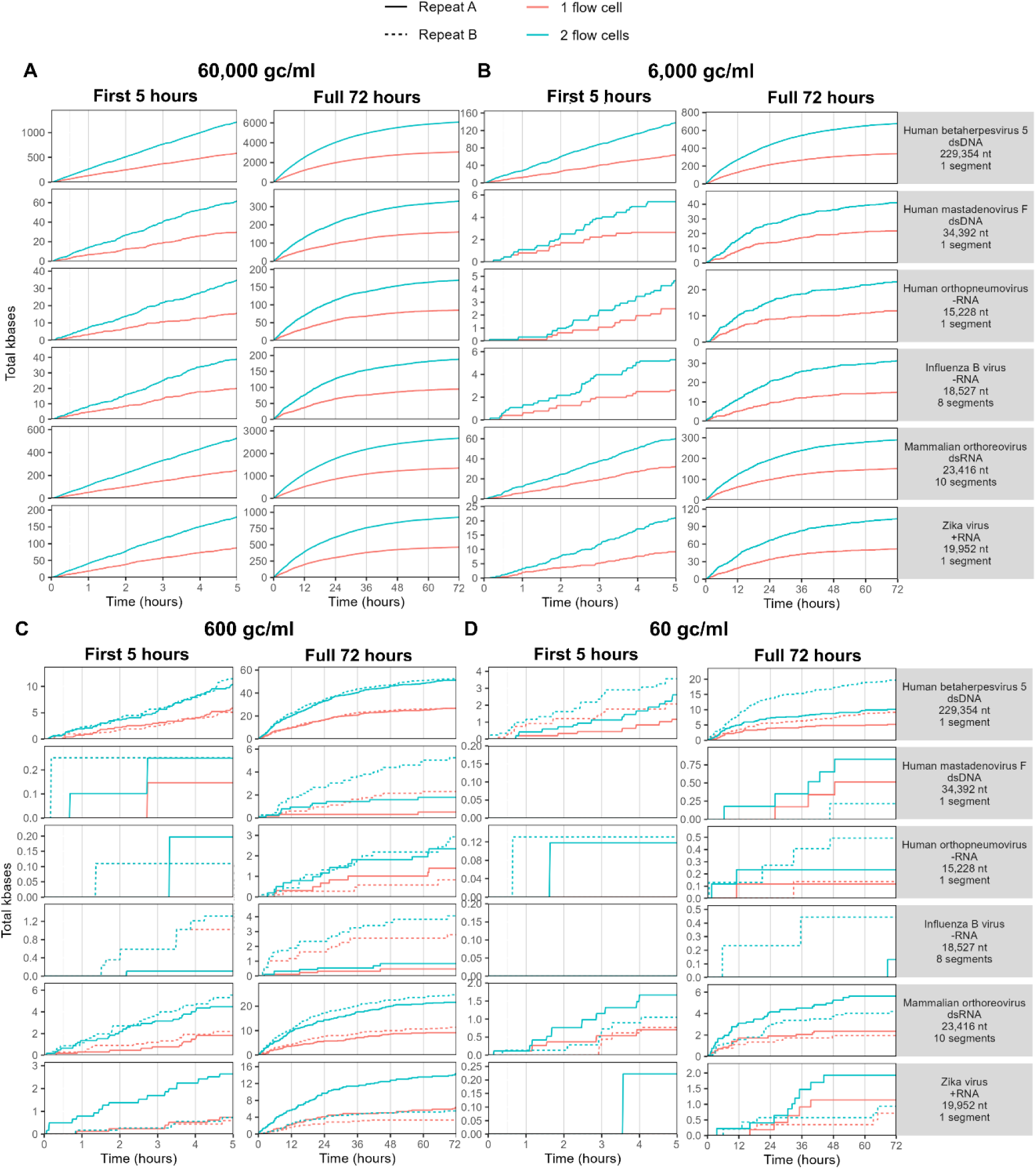
Viral detection over time. Detection of mock community viruses with the ONT + Twist CVRP over time, using reads classified with EPI2ME, measured by kbases of sequenced reads aligning to reference genomes. Times refer to when the read was sequenced. Eight samples were multiplexed per flow cell. Data from two combined flow cells are shown in blue. For 600 gc/ml and 60 gc/ml, replicates are shown with a dotted line. A: 60,000 gc/ml. B: 6,000 gc/ml C: 600 gc/ml. D: 60 gc/ml

**Figure 3:**
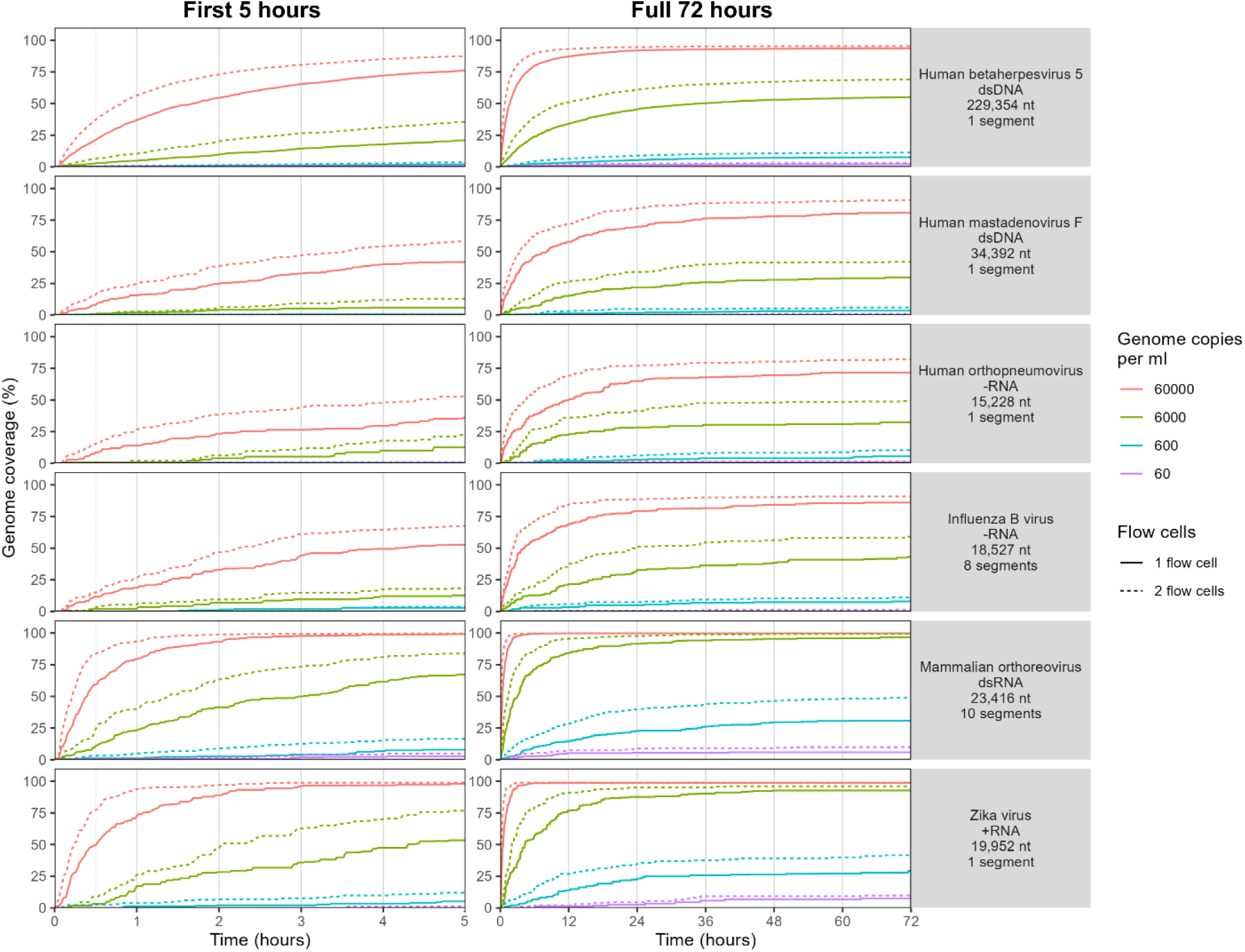
Viral genome coverage over time. Genome coverage of mock community viruses with the ONT + Twist CVRP over time, using reads classified with EPI2ME. Times refer to when the read was sequenced. Eight samples were multiplexed per flow cell. Colours refer to viral concentrations. Dotted lines show the combined data from two flow cells. Results are the average of two replicates for 600 gc/ml and 60 gc/ml.

Doubling the sequencing depths by using a second PromethION flow cell allowed detection of all six viruses by the end of the sequencing run at 72 hours. Additionally, three additional viruses at 600 gc/ml and two at 60 gc/ml were detected within five hours in at least one replicate (**Figure 2C&D**). It also allowed the detection of two additional viruses at 60 gc/ml in at least one replicate at 72 hours (**Figure 2D**). At five hours, the use of two flow cells enabled increases in genome coverage of up to 23.2% compared to one flow cell, for Zika virus at 6,000 gc/ml (**Figure 3**). Sensitivity at 60 gc/ml may be reduced when data from the negative controls is considered – see sensitivity and specificity section.

#### Comparison with untargeted ONT

We then compared the results with untargeted ONT sequencing data previously published on the same mock community samples (**Figure 1C)** [17]. The data volume was normalized as if eight samples were sequenced (including DNA and RNA seq for untargeted ONT) on a single flow cell for each method (**Table S4**). For this analysis, data was aligned to the reference genome sequences of the viruses in the mock community rather than using a specific classifier such as EPI2ME (see next section). The targeted approach provided substantial increases in sensitivity and genome coverage compared to untargeted ONT, identifying all six viruses at all concentrations compared to five, three, two and none for untargeted ONT at 60,000, 6,000, 600 and 60 gc/ml respectively (**Figure 4A, Table S5**). Genome coverage was improved for all viruses at all concentrations, with an average increase in percentage genome coverage of 45.4%, from 44.6% to 90.0%, at 60,000 gc/ml and 52.3%, from 9.84% to 62.2%, at 6,000 gc/ml, where viruses were detected by both approaches (**Figure 4B**). PCR duplicate rates were lower than untargeted ONT at 60,000 gc/ml and lower than previously published duplicate rates for the Twist CVRP (**Figure S2**) [17].

**Figure 4:**
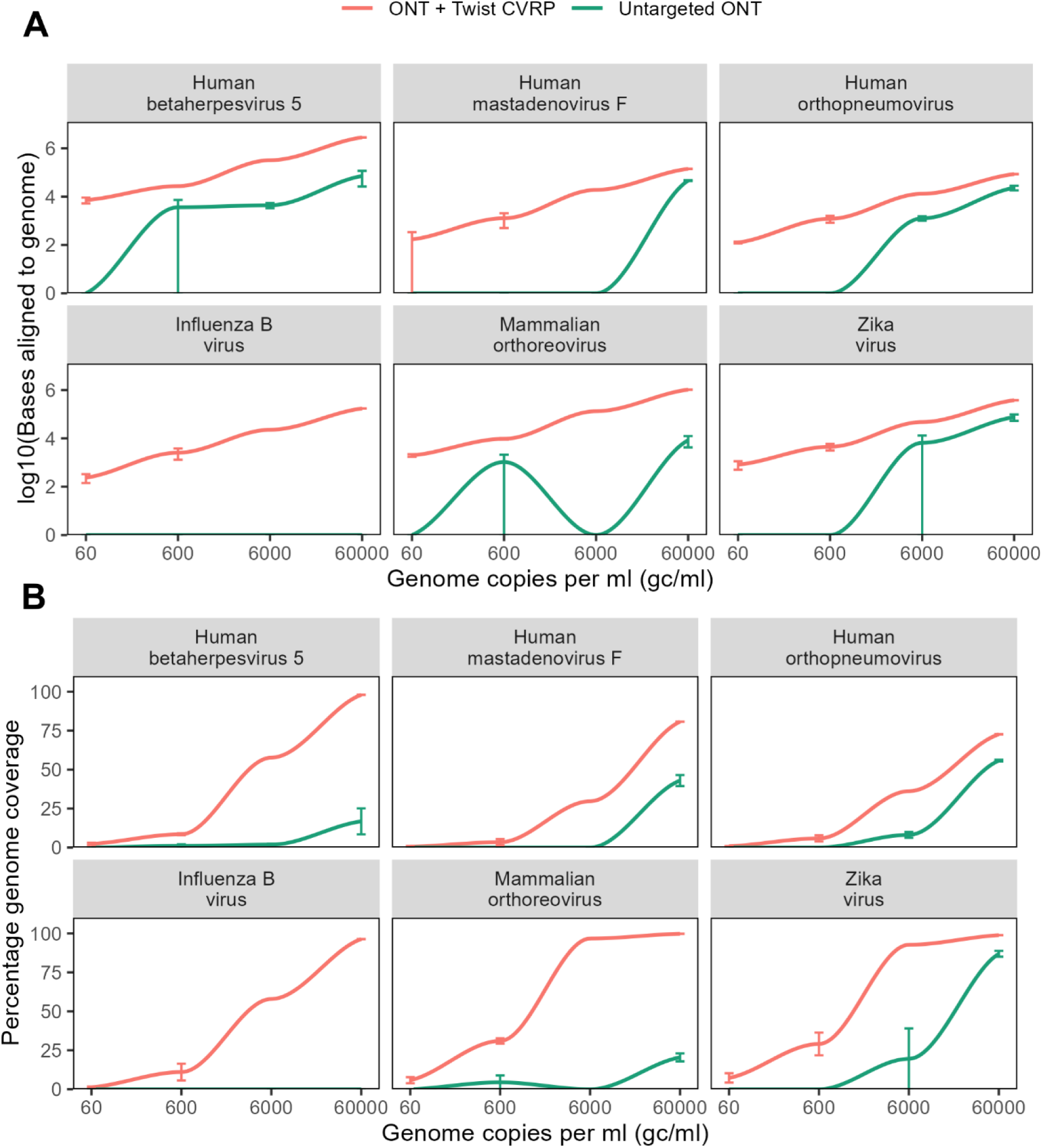
Comparison of ONT + Twist CVRP with untargeted ONT. Normalised comparison of viral detection and genome coverage between untargeted sequencing and the Twist CVRP, both sequenced with ONT. Error bars show range between technical replicates (no replicates performed for ONT + Twist CVRP at 60,000 gc/ml and 6000 gc/ml). A: Log10 of bases aligned to reference genome. B: Percentage reference genome coverage.

In the targeted data, two viruses, human betaherpesvirus 5 and mammalian orthoreovirus, were detected in one or more of the negative control samples, at levels of 13 reads or fewer (**Table S3**). 90% of these reads contained the expected index sequences, suggesting that they represented contamination prior to indexing. 10% (two reads) did not contain both the expected index sequences, suggesting there may have been an error in demultiplexing. This was at lower levels than found for those viruses in the mock community samples, as demonstrated by the sensitivity analysis below (**Table S3**). Viruses were not detected in the negative controls for the untargeted ONT data, possibly due to the lower overall sensitivity of this method.

#### Sensitivity and specificity

A range of bioinformatics tools were used to evaluate the sensitivity and specificity of viral detection. We tested EPI2ME [43], ONT’s recommended analysis tool which can be run in real time, Kraken2 [35] and Bracken [36] run through the nf-core taxprofiler pipeline [34], and CZID [38], a freely available online tool for metagenomics analysis. To reduce the number of false positive species identified, previously determined thresholds based on abundance and comparison with the negative control [17] were applied to the output. The sensitivity of the tools before application of thresholds was similar, with all tools able to detect all viruses at 600-60,000 gc/ml and providing sensitivities of 0.75-0.85 at 60 gc/ml (**Figure 5A**).

**Figure 5:**
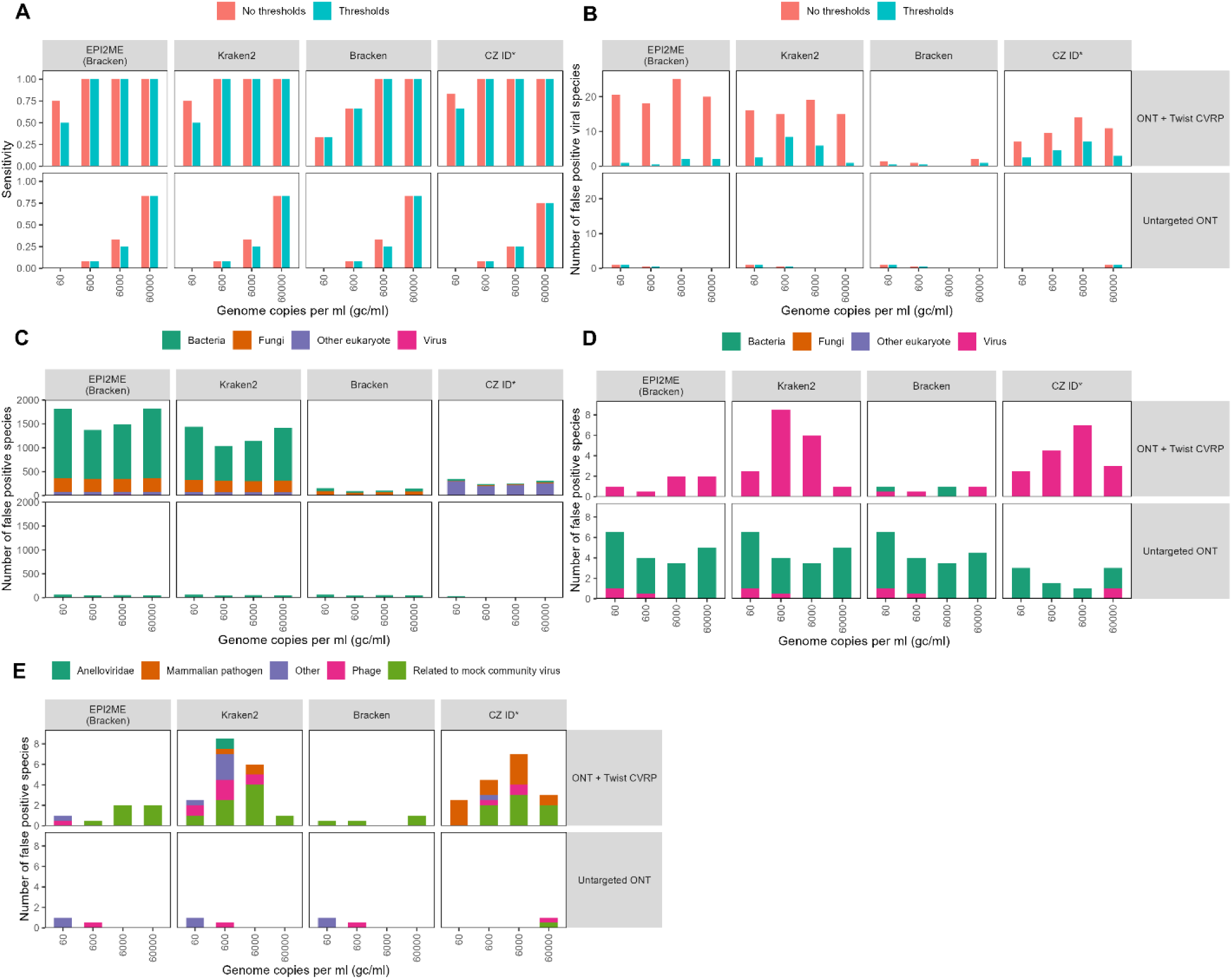
Taxonomic classification and false positive rates. A: Sensitivity of detection of six viral species in mock community before and after thresholds are applied. B-D: Number of false positive species (species not present in mock community). B: False positive viral species before and after thresholds are applied. C: Total false positive species before thresholds. D: Total false positive species after thresholds. E: Classification of false positive viral species that remained after application of thresholds. Values shown are the average of two technical replicates, except ONT + Twist CVRP at 60,000 gc/ml and 6000 gc/ml.

Before the application of thresholds, a higher number of false positive species (defined as species not present in the mock community) were detected with ONT + Twist CVRP compared to the untargeted approach (**Figure 5B&C**). This may be due to the overall higher sensitivity of the targeted approach combined with the shorter length of the reads (a mean N50 of 160 bp compared to 3.5 kb for untargeted ONT) resulting in greater bioinformatic misclassification. After application of thresholds, there were fewer than 10 false positive species in all samples (**Figure 5D**). There were more viral false positives for the Twist CVRP compared to untargeted ONT, reflecting the fact that the panel targets viruses only (**Figure 5D**). The false positive viruses were mostly not clinically relevant (**Figure 5E**). Some Anelloviridae species such as TTV were identified, probably due to contamination of the human genetic material and the difficulty of assigning reads to a specific species meaning that these were not all removed by comparison to the negative control (**Figure 5E**). Some species closely related to members of the mock community were also identified (**Figure 5E**). A small number of false positive mammalian pathogens were identified, particularly by CZID (**Figure 5E**).

#### Effect of read lengths, basecalling, demultiplexing and trimming on data output and sensitivity

In this study, raw read lengths were around 450 bp (**Table 1**), since we adapted an existing short read protocol without additional modifications. This is much shorter than standard ONT reads with the rapid PCR barcoding kit, which may be 2-3 kb in length (**Table S4**). To increase cost and time efficiency, multiple samples are pooled before hybridization and sequenced on the same flow cell. To demultiplex samples after sequencing, unique dual identifiers (UDIs) are used. The index at the beginning and end of the read must be correctly identified by demultiplexing tools to assign a read to a sample. Accurate demultiplexing is necessary to reduce false positive species detections arising from incorrectly assigned reads.

When the data was demultiplexed using UDIs, 67.5% of the reads could not be assigned to a sample and were excluded from further analysis. This was higher than the proportion of unassigned reads (48.3%) for untargeted ONT sequencing, possibly because the index sequences for the targeted approach are shorter than standard ONT barcodes. Basecalling with the super high accuracy (SUP) model, which is not available for real-time analysis, led to a 19.0% increase in reads successfully assigned to barcodes compared to basecalling with the high accuracy (HAC) model (**Figure S1B**). This was the result of both an increase in the total number of reads that passed the quality filters and a higher proportion of SUP reads being assigned to a sample, presumably because of better quality and more intact index sequences (**Table S1**).

We used dual-end barcoding for demultiplexing to ensure specificity of viral detection within each sample (**Figure S1**). However, up to 70% additional bases may be obtained per sample using single-end demultiplexing (**Figure S1**). Once a virus is confidently identified using dual end reads, additional reads that only passed single end demultiplexing could be used to obtain further genome data. Furthermore, future technical improvements could allow super-high accuracy (SUP) basecalling to be performed in real time, further increasing the number of reads available for analysis. Using single end demultiplexing with SUP resulted in a mean increase of 32.7% of bases aligned to the mock community virus genomes at 60,000 gc/ml (**Table S6**). This illustrates that advances in basecalling and demultiplexing could further increase the sensitivity of this method.

The presence of ONT, Illumina and Twist adaptor sequences in the reads, together with the relatively short total read length (N50 = ∼450 bp), meant that around two thirds (300 bp) of every read consisted of adaptors and other technical sequences rather than biologically relevant data (**Figure S1A**). This, together with the low efficiency of demultiplexing, meant that although a total of 26.8 Gb bases were obtained following HAC basecalling, only 9.74 Gb (36.3%) remained following demultiplexing and 3.38 Gb following trimming, meaning that only 12.6% of the total sequence data was biologically relevant (**Table 1**). Increasing this proportion could increase the sensitivity and speed of viral detection without raising sequencing costs.

### Whole genome sequencing

The conversion method is not limited to use with the Twist CVRP and it can be used to convert any capture-based method for use with the ONT rapid barcoding kit. To demonstrate this, we sequenced two mock samples with the Agilent SureSelect XT protocol and ONT on a MinION flow cell. We obtained 0.89-1.01 million reads and 0.22-0.26 Gb per sample, with an N50 of 523-540 bp before and 325-336 bp after trimming (**Table 2**). 49.1-50.0% of each read was lost during trimming, a lower proportion than for the Twist protocol, where roughly two thirds of each read was lost **(Table 1).** This is because the raw reads were longer with the SureSelect method **(Table 2)**. At the end of the run, we obtained >99.8% genome coverage of at least 1X for human betaherpesvirus 5 and SARS-CoV-2, with average sequencing depths ranging from 471-535X (**Figure 6**).

**Figure 6:**
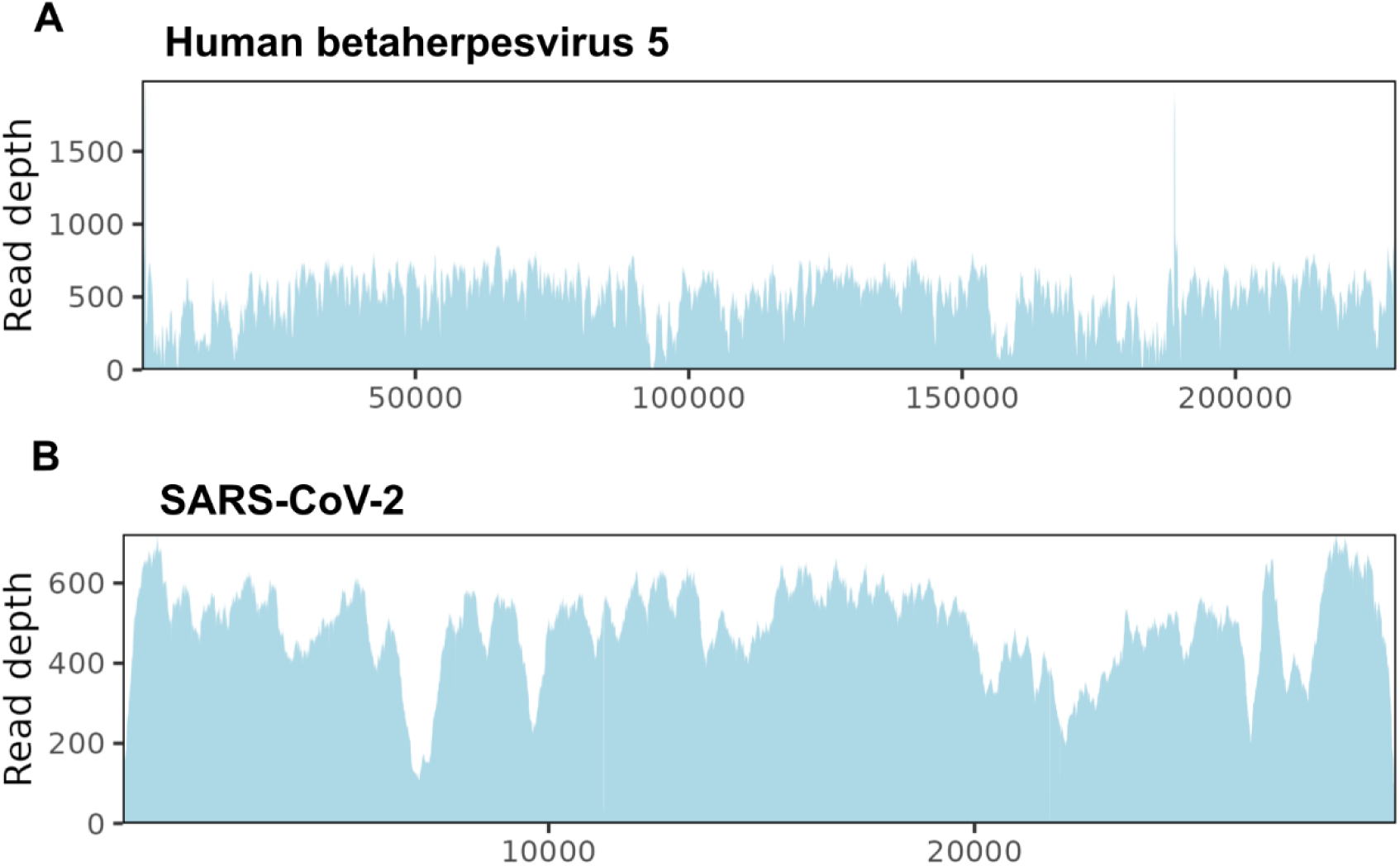
Whole genome sequencing with Agilent SureSelect + ONT. Reference genome coverage of human betaherpesvirus 5 (A) and SARS-CoV-2 (B) from whole genome sequencing with the SureSelect protocol and ONT.

**Table 2:**
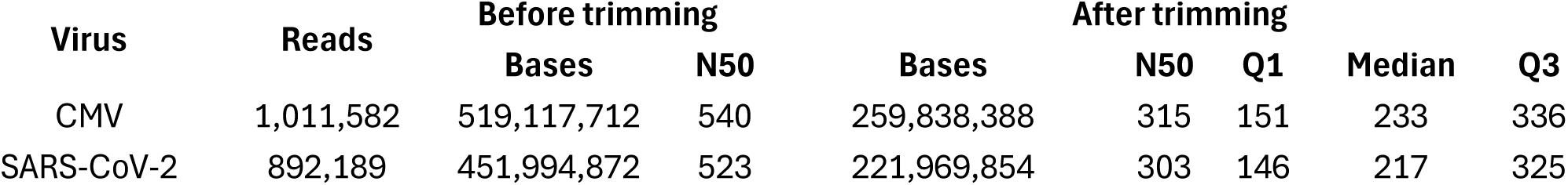
Sequencing output for Agilent SureSelect.

| Virus | Reads | Before trimming |  | After trimming |  |  |  |  |
| --- | --- | --- | --- | --- | --- | --- | --- | --- |
|  |  | Bases | N50 | Bases | N50 | Q1 | Median | Q3 |
| CMV | 1,011,582 | 519,117,712 | 540 | 259,838,388 | 315 | 151 | 233 | 336 |
| SARS-CoV-2 | 892,189 | 451,994,872 | 523 | 221,969,854 | 303 | 146 | 217 | 325 |

## Discussion

In this study we developed a fast universal method that allows the rapid implementation of ONT with existing hybridization capture methods, without significant increases in turnaround times and costs. While untargeted ONT approaches allow the detection of highly abundant viruses, sensitivity for low viral loads is limited, particularly for RNA viruses [15,17,44,45]. By combining ONT with the TWIST CVRP, a target capture panel for over 3000 viruses, we increased the sensitivity of ONT metagenomics by 10 to 100-fold compared to untargeted approaches, enabling the detection of low viral loads **(Figure 4).** This increase in sensitivity is similar to that observed for hybridization capture workflows for short-read sequencing [17,45]. Viral genome coverage was also increased compared to untargeted approaches **(Figure 4)**, which could facilitate applications such as phylogenetic and epidemiological analysis. The target capture method overcomes some limitations of alternative protocols for increasing the sensitivity of ONT, such as differential lysis followed by nuclease digestion of human nucleic acids, which may fail to detect certain pathogens due to loss of cell-free DNA and lysis biases. In addition, retaining host material could allow the simultaneous detection of host biomarkers without the need for additional samples or laboratory protocols, as part of a precision medicine approach.

Real-time analysis allowed the detection of all viruses at high viral loads (60,000 and 6000 gc/ml) within two hours of sequencing, providing actionable results early in the run (**Figure 2**). Longer runs of up to 72 hours were required to detect low viral loads (60 gc/ml) (**Figure 2**). Reducing the number of samples pooled per hybridization reaction can improve the sensitivity of early reports and reduce the need for longer runs **(Figure 3)**, although this will lead to a cost increase per sample. Access to preliminary results during the sequencing run can lead to reduced turnaround times especially for high viral load samples. Given that the duration of the Twist CVRP hybridization capture protocol is approximately 1.5 days, early reports can provide actionable data within the second day of the workflow **(Figure 1B**). Real-time reports have been incorporated in rapid ONT metagenomic workflows, providing preliminary results just after 30 min and 2 hours of the start of the sequencing run [15]. Additional modifications to the current hybridization capture workflow could lead to reductions in turnaround times and increased sensitivity. Current methods often require long hybridizations, such as overnight in the case of the Twist CVRP, which is a significant contributor to their overall turnaround times. Use of fast-hybridization alternatives could lead to significant improvements in sample processing times. However, it will be necessary to validate that these reductions do not impact sensitivity.

The reads obtained in this analysis averaged 160 bp after adaptor trimming, which is comparable to those produced by short read technologies. Raw reads were short (around 450 bp) since the short-read hybridization capture protocol was not modified and adaptor trimming led to the loss of around 300 bp per read. However, a key advantage of ONT is its ability to produce longer reads, averaging several kb in length, which can aid genome assembly and increase specificity of taxonomic classification. In future, libraries could be constructed with longer inserts, generating greater read lengths with an increased proportion of usable sequence data compared to adaptor regions. However, longer reads will require longer extension times during both pre- and post-capture PCR steps, leading to a potential increase in turnaround times.

Pooling of samples for hybridization makes library preparation more cost effective. During sample demultiplexing, a lower proportion of reads was successfully assigned to a sample for ONT + Twist CVRP compared to untargeted ONT, possibly due to the shorter index sequences for Twist compared to the ONT barcodes. Increasing the length of the Twist barcodes could enable more efficient demultiplexing, increasing the volume of usable sequencing data. Use of more accurate basecalling models such as SUP could also increase data volumes (**Figure S1**). While the use of real-time SUP is currently limited to high spec GPUs and PromethiON 24/48, recent improvements in basecalling have led to significant speed increases and further developments could allow it to be performed in real time on a wider variety of machines.

While this fast targeted method using ONT sequencing has clear applications in diagnostic metagenomics, especially in clinical situations where sensitivity is key, targeted methods have limitations. Efficient detection of pathogens depends on the breadth and quality of the probe sets and further work is needed to optimize these, for example by including bacteria, fungi and parasites, while minimizing the costs of large panels. Targeted metagenomics may also miss emerging pathogens of clinical interest, especially where these are more than 20% divergent from existing probes [21,22]. Further work is required to establish rates of detection of non-targeted species in clinical samples.

Our conversion method is not specific to targeted metagenomics and could in theory allow use of ONT with any hybridization capture method designed for short read sequencing. As proof of concept, we successfully converted SureSelect XT libraries for use with ONT and generated whole genome sequences of human betaherpesvirus 5 and SARS-CoV-2. (**Figure 6**). SureSelect can also be used with panels targeting larger numbers of pathogens and could be used in targeted metagenomics applications [46]. In whole genome sequencing applications, minimizing the turnaround time of the PCR step is less critical, meaning that protocols could be adapted to generate longer fragments (>1 kb). This may be beneficial for the study and characterization of viruses with long repeat regions and high frequency of recombination events [47–49].

## Conclusions

This method combines the speed and low costs of ONT with the increased sensitivity of hybridization capture approaches. Here, we demonstrate its utility in improving the sensitivity and turnaround times of viral metagenomics. In future, it could be used not only in pathogen sequencing, but in a wide variety of clinical contexts, such as exome panels to identify structural variation, pharmacogenomics and cancer panels.

## Data Availability

The raw data produced in this study is available in the European Nucleotide Archive (ENA) with accession PRJEB87418 (data will be made available after publication).

## List of abbreviations

Twist CVRP: Twist Comprehensive Viral Research Panel
ONT: Oxford Nanopore Technologies

## Declarations

### Ethics approval and consent to participate

There were no human participants in this study. The human nucleic acids used were commercially available.

### Consent for publication

Not applicable

### Availability of data and materials

The raw data produced in this study is available in the European Nucleotide Archive (ENA) with accession PRJEB87418 (*data will be made available after publication)*.

### Authors’ contributions

OETM and SB devised the method. OETM, SB, RW, SM and JB designed the study. OETM, LF, TB, and HT performed laboratory work. SB analysed the data. OETM, SB, SM and JB wrote and revised the manuscript. PS, HH, TG and RW revised the manuscript. All authors read and approved the final manuscript.

## Acknowledgements

Not applicable

---

**Figure S1:**
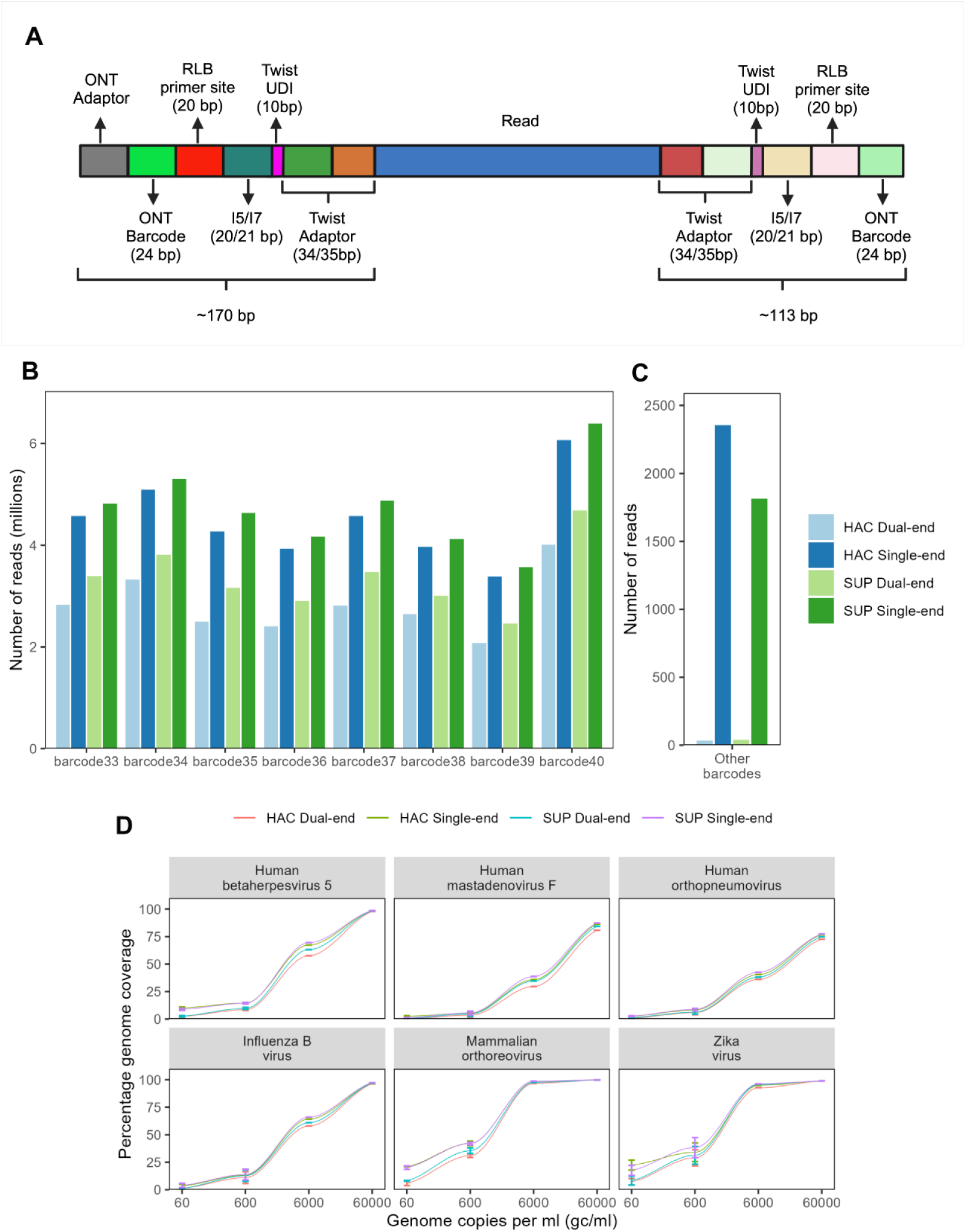
Basecalling and demultiplexing. A: Structure of read produced by Twist CVRP and conversion using four-primer approach. B: Number of reads after demultiplexing with high accuracy (HAC) and super high accuracy (SUP) basecalling and dual and single end demultiplexing using dorado. C: Number of reads assigned to barcodes not expected to be present in the sample, suggesting they were incorrectly assigned. D: Genome coverage of viruses with different basecalling and demultiplexing approaches.

**Figure S2:**
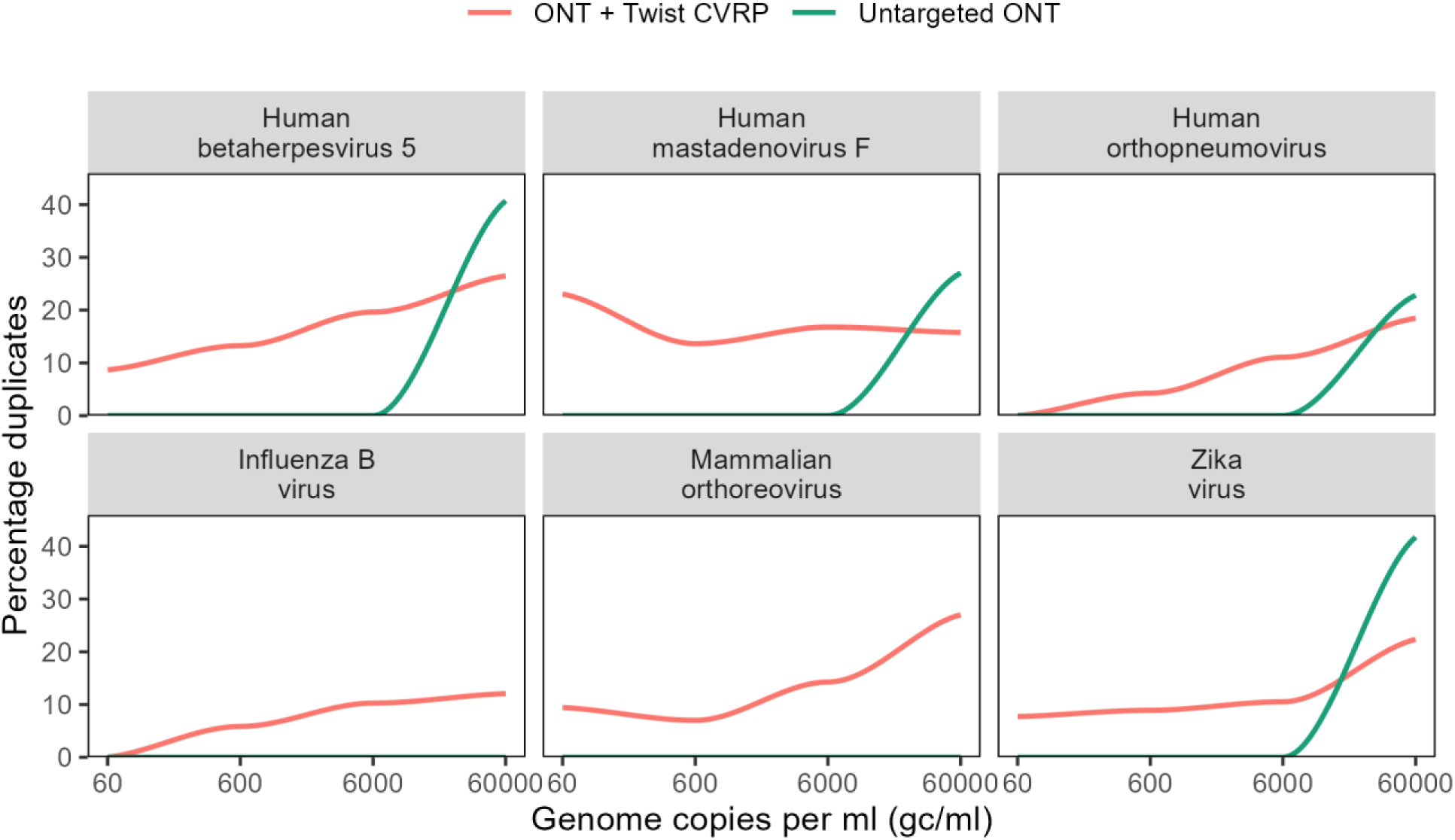
PCR duplicate rates. Percentage of reads aligning to the viral reference genomes that were PCR duplicates for the ONT + Twist CVRP and untargeted ONT. Duplicate rates are shown as 0 where there were no assigned reads at that viral concentration.

## Supplementary Tables

Table S1: Reads passing basecalling quality filters and demultiplexing with HAC and SUP

Table S2: Sequencing output at 72 hours with one and two flow cells

Table S3: Results of viral genome alignment for ONT + Twist CVRP at 72 hours

Table S4: Sequencing data volumes for untargeted ONT (this data was generated in a previous study [17])

Table S5: Viruses detected using alignment by targeted and untargeted ONT

Table S6: Increase in bases assigned with SUP and single-end demultiplexing compared to HAC with dual-end demultiplexing

